# Global approaches to postmortem genetic testing after sudden cardiac death in the young: A survey among healthcare professionals

**DOI:** 10.1101/2020.12.23.20248816

**Authors:** Lieke M. van den Heuvel, Judy Do, Laura Yeates, Heather MacLeod, Cynthia A. James, Johan Duflou, Jonathan R. Skinner, Christopher Semsarian, J. Peter van Tintelen, Jodie Ingles

**Author notes:** **Address for correspondence:** Associate Professor Jodie Ingles | Cardio Genomics Program | Centenary Institute, Locked Bag 6 Newtown NSW, 2042 Sydney Australia | P. 02 9665 6100 | E. | Twitter. @jodieingles27. **CONFLICT OF INTEREST NOTIFICATION** L.M. van den Heuvel: No disclosures. J. Do: No disclosures. L. Yeates: No disclosures. H. MacLeod: No disclosures. C.A. James: Research grant support from Boston Scientific Corp. J. Duflou: No disclosures. J.R. Skinner: No disclosures. C. Semsarian: No disclosures. J.P. van Tintelen: No disclosures. J. Ingles: Research grant support from Myokardia, Inc.

## Abstract

**Purpose:** Thorough investigation of sudden cardiac death (SCD) in those aged 1-40 years commonly reveals a heritable cause, yet access to postmortem genetic testing is variable. We explore practices of postmortem genetic testing and attitudes of healthcare professionals worldwide.

**Methods:** A survey was administered among healthcare professionals recruited through professional associations, social media and networks of researchers. Topics included practices around postmortem genetic testing, level of confidence in healthcare professionals’ ability, and attitudes towards postmortem genetic testing practices.

**Results:** There were 112 respondents, with 93% from North America, Europe and Australia and 7% from South America, Asia and Africa. Only 30% reported autopsy as mandatory, and overall practices were largely case-by-case and not standardised. North American respondents (87%) more often perceived practices as ineffective compared to those from Europe (58%) and Australia/New Zealand (48%, *p=*0.002). Where a heritable cause is suspected, 69% considered postmortem genetic testing and 61% offered genetic counseling to surviving family members; financial resources varied widely. Half believed practices in their countries perpetuated health inequalities.

**Conclusion:** Postmortem genetic testing is not consistently available in the investigation of young SCD despite being a recommendation in international guidelines. Access to postmortem genetic testing, guided by well-resourced multidisciplinary teams, is critical in ascertaining a cause of death in many cases.

## INTRODUCTION

Sudden cardiac death (SCD) in the young is a rare but tragic event and the impact on the family and the community is significant. Postmortem pathological examination may identify structural changes. In older patients, the majority of SCD can be attributed to coronary artery disease (1). Cardiomyopathies, potentially genetic, may be another cause of SCD identified at postmortem examination (2). However, in 30-40% of SCD in young people (<35-40 years), no structural changes are identified at postmortem examination (3, 4). While SCD in a young family member is devastating, not knowing the cause of death can limit clinical and genetic screening options adding to the uncertainty (5). Failing to identify a heritable cause of death results in failure to identify other family members who may be at risk of SCD.

In cases where no structural abnormalities are identified at postmortem evaluation and an underlying inherited arrhythmia syndrome is suspected, genetic testing as part of a multidisciplinary investigation of sudden unexpected death, and this practice is supported by international guideline documents (6-9). Postmortem genetic testing may identify an underlying inherited cardiac disease in 15-40% of cases (3, 10-12). It thereby allows for determination of the risk of family members, that can ultimately lead to prevention of morbidity and mortality in these families. Genetic testing in combination with clinical screening of family members can increase the yield, ranging from 33-53% (13-15). Furthermore, from a psychosocial perspective, identifying a cause of death can give families much needed answers.

Unfortunately, in many countries, autopsy and/or postmortem genetic testing is not routinely performed after unexpected SCD in the young (8, 16). While national and sometimes continental regulations are in place, the rate of postmortem investigation and genetic testing, as well as the procedures for doing this, are vastly different between countries (8). These differences likely reflect different legislative requirements, cultural differences, inequities in funding and healthcare services, models of care allowing access to relatives, and routine assessment of families in specialized clinics (8). There are few data revealing how practices vary worldwide. Here we explore current practices of postmortem genetic testing worldwide, as well as attitudes towards these practices of healthcare professionals involved.

## MATERIALS AND METHODS

### Participants and recruitment

For this cross-sectional survey, healthcare professionals involved in postmortem genetic testing, including cardiologists, genetic counselors, clinical geneticists, and pathologists, were asked to participate. Healthcare professionals were approached through newsletters of relevant societies (both national and international societies) (see Supplementary Material S1), by social media (Twitter) and/or directly by authors JI and PvT. The survey was provided online, using Redcap survey software (17). The survey was anonymous, and completion of the survey was deemed as informed consent given. Ethical approval of the local institutional ethics committee was obtained.

### Data collection

The survey was developed by a psychologist (LvdH) with input from a multidisciplinary team with experience in the field, including genetic counselors, clinical researchers, cardiologists and a forensic pathologist. The survey included closed questions with opportunity to provide open text additional information. Questions addressed the following topics: (1) Practices around postmortem genetic testing in their clinic/country, (2) Level of confidence in their ability to be involved in practices around postmortem genetic testing in their clinic/country, (3) Attitudes towards postmortem genetic testing practices in their country/clinic. Demographics collected included country, profession, and years of experience. The survey is included as Supplementary Material (Supplementary Material S2).

### Data analysis

Quantitative data analysis was conducted in SPSS (Version 25). Data were exported from Redcap. Descriptive statistics were used to describe the study sample and the survey responses. To assess differences in responses between regions of residence (comparing practices and attitudes Europe, Australia/New Zealand and North America), Chi square analyses were performed to compare groups. Due to low numbers of responses in the other continents (i.e., Asia, South America and Africa) only descriptive data are presented. A Bonferroni corrected *p* value of <0.05 was considered significant. Open answers were coded by LH and LY and a codebook was created. Differences in codes were discussed by LH and LY until agreement was met. Quotes from open answers are included to illustrate the results.

## RESULTS

### Demographics

In total, 112 respondents participated in the survey. Figure 1 shows the sociodemographic characteristics of participants. Most participants were cardiologists (N=41, 37%) or genetic counselors (N=36, 33%). Other professions included clinical geneticists, pathologists, scientists, cardiac nurses, and cardiology fellows. Most participants were from countries in Europe (N=45, 40%), Australia/New Zealand (N=33, 30%), and North America (N=26, 23%). The remainder (N=8, 7%) came from South America, Asia and Africa. The majority of participants worked in a specialized cardiogenetics clinic (N=64, 58%). Others worked in a general cardiac (N=15, 14%) or genetics clinic (N=16, 14%) or forensic medicine (N=8, 7%). The remainder worked in pathology, pediatric electrophysiology or a medical research institute (N=7, 7%). Participants had between 2-20 years of experience in their profession, and in working with families with SCD in the young. There was an even mix of participants in terms of their overall number of SCD cases they had been involved with in their career to date, ranging from 1-5 cases (N=13, 13%) to 100-500 cases (N=17, 17%) Two participants had been involved in over 500 sudden death cases during their career (2%).

**Figure 1:**
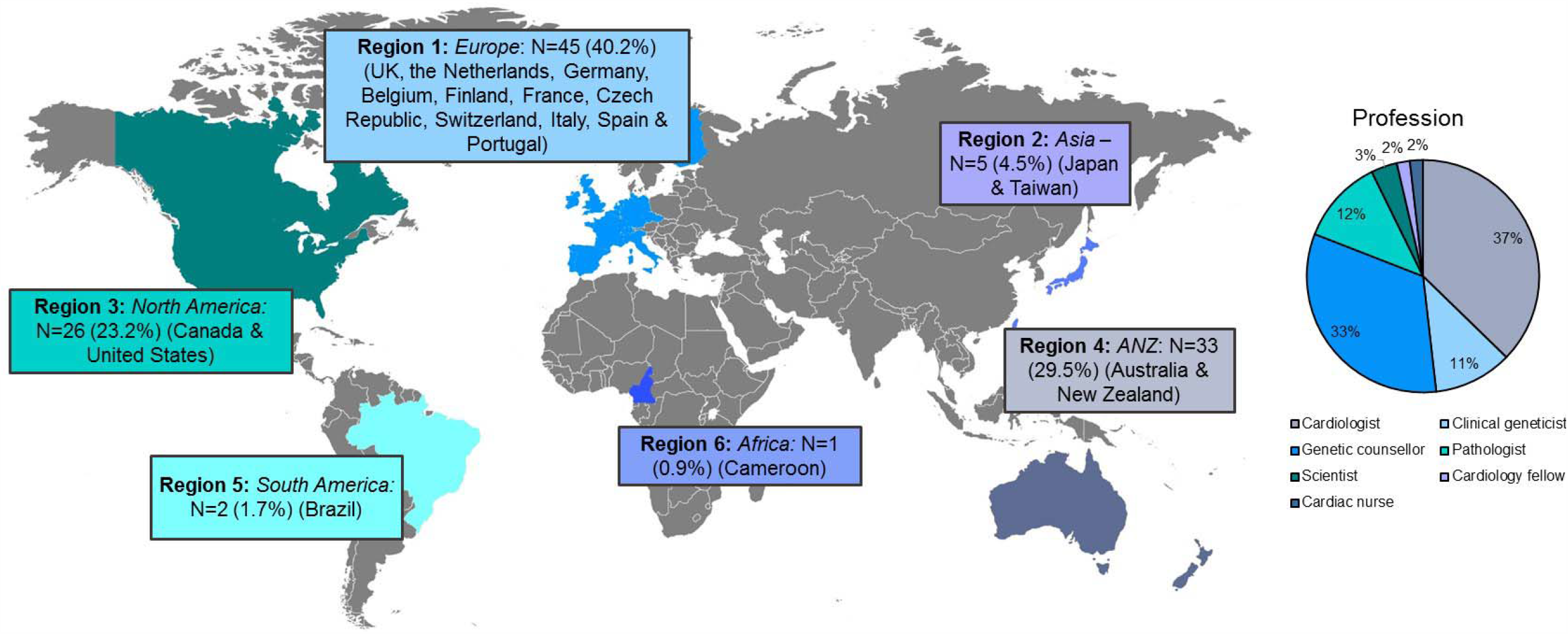
Participants per continent. Respondents per continent and by expertise are shown. Map from vemaps.com.

As shown in Figure 2, almost all participants felt generally confident or very confident in their work activities in sudden unexpected death cases. For 65% of participants (N=64) for whom this question was applicable, they reported that they had not received any specific training on the process of postmortem genetic testing.

**Figure 2:**
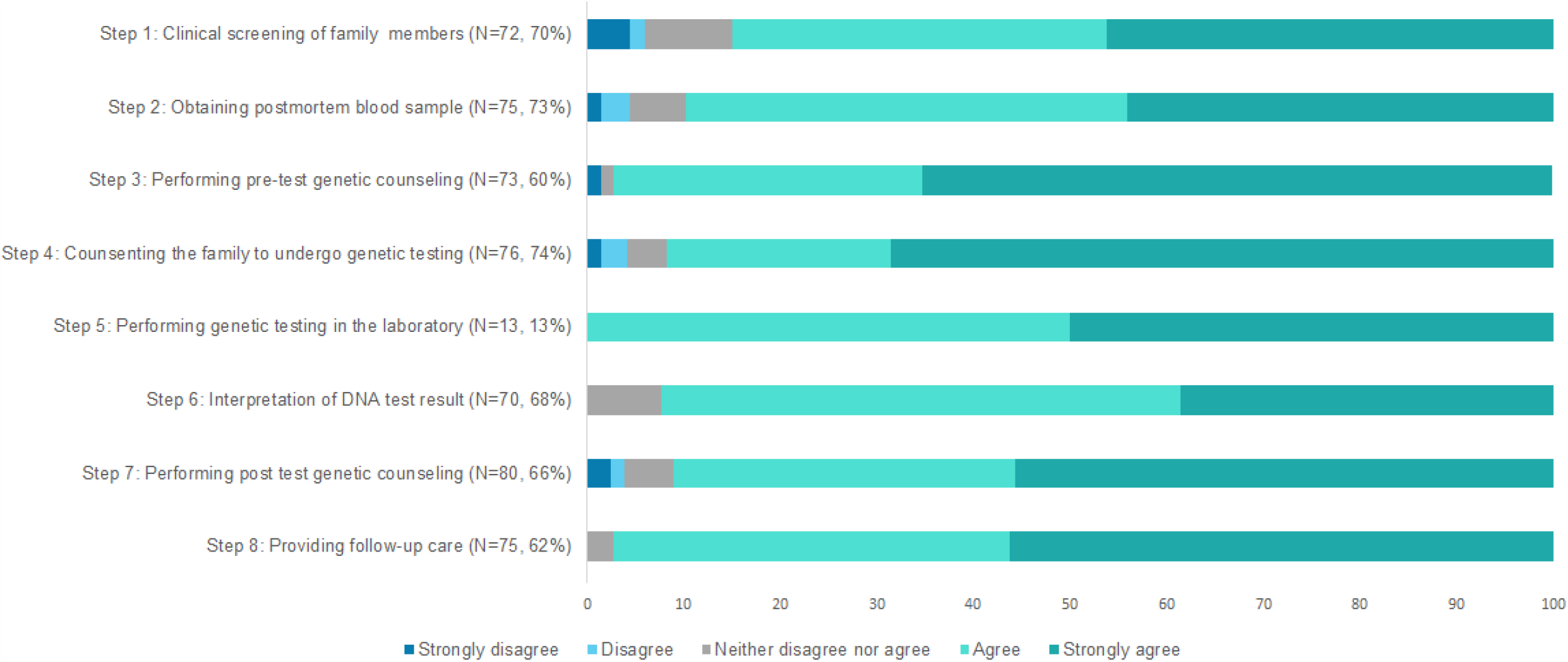
Confidence of respondents in aspects of sudden cardiac death (SCD) care they are involved in. Shown are the percentage of respondents who are involved in each step of the postmortem investigation process, and their overall confidence with this aspect of care. *Presentation of data regarding confidence of participants in the steps participants indicated to be involved in. N shows the number of participants who indicated to be involved in each step. Confidence on a 5 point Likert scale (strongly disagree-strongly agree) is reported in percentages*.

### Guidelines and legal obligations regarding postmortem genetic testing

Table 1 shows the survey responses. With regard to guideline use for the investigation of sudden unexpected death in a young person, there were 38% (N=39) who responded that international or national guidelines are implemented, while 25% (N=26) of participants reported this was either not occurring or they were unsure (N=38, 37%). There was a difference observed between practices in Europe, Australia/New Zealand and North America (*p*=0.002), with participants from the United States and Canada less likely to state that autopsy guidelines were implemented (17%) or that they were unsure (44%), compared to practices in Europe (59%) and Australia/New Zealand (36%) who more often had guidelines implemented.

**Table 1.** Healthcare practices regarding postmortem genetic testing after sudden death in the young

|  | Total | Europe | Australia & New Zealand | North America |  | Other |
| --- | --- | --- | --- | --- | --- | --- |
|  | N (%) | N (%) | N (%) | N (%) | *P* value<sup>b</sup> | N (%) |
| **Guidelines implemented** |  |  |  |  |  |  |
| Yes | **39 (37.9)** | 24 (58.5) | 11 (35.5) | 4 (17.4) | 0.002<sup>c</sup> | 0 (0.0) |
| No | **26 (25.2)** | 8 (19.5) | 3 (9.7) | 9 (39.1) |  | 6 (75.0) |
| Unsure | **38 (36.9)** | 9 (22.0) | 17 (54.8) | 10 (43.5) |  | 2 (25.0) |
| **Autopsy mandatory** |  |  |  |  |  |  |
| Yes | **24 (23.3)** | 9 (22.0) | 8 (25.8) | 6 (26.1) | 0.024 | 1 (12.5) |
| No | **31 (30.1)** | 17 (41.5) | 3 (9.7) | 7 (30.4) |  | 4 (50.0) |
| Unsure | **22 (21.4)** | 4 (9.8) | 13 (41.9) | 5 (21.7) |  | 0 (0.0) |
| Other | **26 (25.2)** | 11 (26.8) | 7 (22.6) | 5 (21.7) |  | 3 (37.5) |
| **Postmortem genetic testing considered** ☐ |  |  |  |  |  |  |
| In case of SD with identified (genetic) cause at autopsy | **71 (68.9)** | 32 (71.1) | 21 (63.6) | 17 (65.4) | NA | 1 (12.5) |
| In case of SUD | **66 (64.1)** | 29 (64.4) | 17 (51.5) | 18 (69.2) |  | 2 (25.0) |
| In case of SUD with ambiguous autopsy findings | **46 (38.0)** | 18 (40.0) | 11 (33.3) | 15 (57.7) |  | 2 (25.0) |
| That depends on the case | **30 (24.8)** | 10 (22.2) | 11 (33.3) | 5 (19.2) |  | 4 (50.0) |
| **Consent of family members required** |  |  |  |  |  |  |
| Yes | **75 (72.8)** | 34 (82.9) | 22 (71.0) | 15 (65.2) | 0.488 | 4 (50.0) |
| No | **10 (9.7)** | 3 (7.3) | 2 (6.5) | 4 (17.4) |  | 1 (12.5) |
| Unsure | **12 (11.7)** | 2 (4.9) | 5 (16.1) | 2 (8.7) |  | 3 (37.5) |
| Other | **6 (5.8)** | 2 (4.9) | 2 (6.5) | 2 (8.7) |  | 0 (0.0) |
| **Blood sample taken** |  |  |  |  |  |  |
| Yes | **26 (25.2)** | 10 (24.4) | 11 (35.5) | 4 (17.4) | 0.002<sup>c</sup> | 1 (12.5) |
| No | **8 (7.8)** | 4 (9.8) | 0 (0.0) | 2 (8.7) |  | 2 (25.0) |
| On a case-by-case basis | **49 (47.6)** | 21 (51.2) | 12 (38.7) | 12 (52.2) |  | 4 (50.0) |
| Unsure | **10 (9.7)** | 1 (2.4) | 8 (25.8) | 0 (0.0) |  | 1 (12.5) |
| Other | **10 (9.7)** | 5 (12.2) | 0 (0.0) | 5 (21.7) |  | 0 (0.0) |
| **Receive autopsy report** |  |  |  |  |  |  |
| Yes | **45 (47.4)** | 18 (51.4) | 16 (53.3) | 10 (43.5) | 0.440 | 1 (14.3) |
| No | **6 (6.3)** | 1 (2.9) | 3 (10.0) | 0 (0.0) |  | 2 (28.6) |
| On a case-by-case basis | **28 (29.5)** | 11 (31.4) | 8 (26.7) | 7 (30.4) |  | 2 (28.6) |
| Other | **16 (16.8)** | 5 (14.3) | 3 (10.0) | 6 (26.1) |  | 2 (28.6) |
| **Genetic counseling provided** |  |  |  |  |  |  |
| Yes | **63 (61.2)** | 25 (61.0) | 23 (74.2) | 13 (56.5) | 0.241 | 2 (25.0) |
| No | **7 (6.8)** | 3 (7.3) | 3 (9.7) | 1 (4.3) |  | 0 (0.0) |
| On a case-by-case basis | **18 (17.5)** | 7 (17.1) | 1 (3.2) | 5 (21.7) |  | 5 (62.5) |
| Unsure | **7 (6.8)** | 4 (9.8) | 2 (6.5) | 0 (0.0) |  | 1 (12.5) |
| Other | **8 (7.8)** | 2 (4.9) | 2 (6.5) | 4 (17.4) |  | 0 (0.0) |
| **Postmortem genetic testing provided** |  |  |  |  |  |  |
| As a clinical test | **49 (48.0)** | 19 (46.3) | 18 (60.0) | 12 (52.2) | 0.924 | 0 (0.0) |
| As a research-based test | **4 (3.9)** | 2 (4.9) | 0 (0.0) | 1 (4.3) |  | 1 (12.5) |
| On a case-by-case basis | **32 (31.4)** | 13 (31.7) | 8 (26.7) | 8 (34.8) |  | 3 (37.5) |
| Not routinely available | **10 (9.8)** | 4 (9.8) | 2 (6.7) | 1 (4.3) |  | 3 (37.5) |
| Other | **7 (6.9)** | 3 (7.3) | 2 (6.7) | 1 (4.3) |  | 1 (12.5) |
| **Postmortem genetic testing may be ordered by** |  |  |  |  |  |  |
| Genetic counselors | **47 (38.8)** | 18 (40.0) | 12 (36.4) | 17 (65.4) | NA | 0 (0.0) |
| Clinical geneticists | **76 (62.8)** | 31 (68.9) | 27 (81.8) | 16 (61.5) |  | 2 (25.0) |
| Cardiologists | **60 (49.6)** | 28 (62.2) | 13 (39.4) | 16 (61.5) |  | 3 (37.5) |
| Pathologists | **22 (18.2)** | 7 (15.6) | 4 (12.1) | 8 (30.8) |  | 3 (37.5) |
| Unsure | **8 (6.6)** | 2 (4.4) | 3 (9.1) | 1 (3.8) |  | 2 (25.0) |
| Other | **14 (11.6)** | 5 (11.1) | 6 (18.2) | 3 (11.5) |  | 0 (0.0) |
| **Postmortem genetic testing funded** |  |  |  |  |  |  |
| By the hospital/healthcare system | **54 (44.6)** | 20 (44.4) | 24 (72.7) | 9 (34.6) | NA | 1 (12.5) |
| Covered by healthcare insurance | **19 (15.7)** | 13 (28.9) | 0 (0.0) | 5 (19.2) |  | 1 (12.5) |
| Family members pay for it themselves | **17 (14.0)** | 4 (8.9) | 3 (9.1) | 8 (30.8) |  | 2 (25.0) |
| It is covered by research funding | **22 (18.2)** | 12 (26.7) | 2 (6.1) | 4 (15.4) |  | 4 (50.0) |
| Other | **19 (15.7)** | 6 (13.3) | 5 (15.2) | 7 (26.9) |  | 1 (12.5) |
| **Clinical screening provided** |  |  |  |  |  |  |
| Yes, concurrent with postmortem genetic testing | **58 (56.3)** | 25 (61.0) | 17 (54.8) | 14 (60.9) | 0.358 | 2 (25.0) |
| Yes, after postmortem genetic testing depending on the result | **23 (22.3)** | 8 (19.5) | 7 (22.6) | 6 (26.1) |  | 2 (25.0) |
| No | **0 (0.0)** | 0 (0.0) | 0 (0.0) | 0 (0.0) |  | 0 (0.0) |
| Unsure | **10 (9.7)** | 5 (12.2) | 1 (3.2) | 0 (0.0) |  | 4 (50.0) |
| Other | **12 (11.7)** | 3 (7.3) | 6 (19.4) | 3 (13.0) |  | 0 (0.0) |
| **Follow-up care family members provided<sup>□</sup>** |  |  |  |  |  |  |
| Yes, clinical screening | **75 (62.0)** | 34 (75.6) | 16 (48.5) | 21 (80.8) | NA | 4 (50.0) |
| Yes, post-test genetic counseling | **78 (64.5)** | 30 (66.7) | 24 (72.7) | 21 (80.8) |  | 3 (37.5) |
| Yes, psychosocial care | **47 (38.8)** | 14 (31.1) | 16 (48.5) | 16 (48.5) |  | 1 (12.5) |
| No | **4 (3.3)** | 0 (0.0) | 0 (0.0) | 0 (0.0) |  | 4 (50.0) |

<sup>□</sup> Statistical significance tests are not performed on questions with multiple answer options
<sup>b</sup>Chi square analyses were performed on continents Europe, Australia/New Zealand and North America, due to low numbers of responses from other continents
<sup>c</sup>Statistically significant, based on a bonferroni-corrected p value of 0.003 (0.05/16) for responses displayed.

### Autopsy

Only 24 (23%) responded that autopsy is mandatory, or that they were unsure about this (N=22, 21%). There were 26 (25%) participants who provided explanations stating that it is only mandatory in cases of a SCD under 18 years of age, that the coroner, medical examiner or pathologist decides, and that there is a difference between provinces/states in their countries. Relatively more respondents from Europe reported that autopsy was not mandatory (42%), compared to Australia/New Zealand (10%) and North-America (30%). Some participants (N=13, 42%) from Australia/New Zealand indicated they were unsure whether autopsy is mandatory in their countries. This difference was however not considered significant using a Bonferroni-corrected significance level (*p=*0.024). Almost half of participants (N=49, 48%) reported that a blood sample was taken on a case by case basis, and 25% (N=26) responded that this was a standard procedure. Participants from Australia/New Zealand more often reported that a sample for genetic testing is taken as standard part of the postmortem investigation (36%), compared to participants from the Europe (24%) and North-America (17%) (*p*=0.002). Where more information was provided, respondents stated it depends on the clinician/pathologist or mortuary involved. For example, one participant reported:

“*It very much depends on the pathologist performing the autopsy. Given the guidelines, it should be taken in every case but I am sure that most clinical pathologists do not bother*.*”* (Europe)

Many participants responded that they receive the autopsy report directly (n=45, 44%), and 28 (27%) reported that this only occurred in certain cases. Sixteen participants gave an open answer, stating that family members visiting the cardiogenetics clinic are asked to bring the report or that they specifically request it from the coroner. A participant explained:

“[*I receive the autopsy report] when the forensic team have referred the family to me, or when the family bring it in themselves if referred by others*.” (Australia)

Of the participants for whom this question was applicable, most (N=73, 81%) said that they use the autopsy report in their clinic.

### Postmortem genetic testing

A majority (N=71, 69%) responded that postmortem genetic testing is considered for SCD where a genetic cause is suspected. In cases of SCD where no cause is found after postmortem investigation, 66 participants (64%) responded that postmortem genetic testing would be indicated. Further, 75 (73%) reported that consent for performing postmortem genetic testing is obtained from surviving relatives, with six participants stating that this depends on whether the coroner or a medical specialist requested postmortem genetic testing.

The majority of participants (N=63, 61%) stated genetic counseling was provided to surviving relatives in their center. Others reported that this was provided on a case-by-case basis, or that this was never provided or were unsure. Eight participants (8%) provided explanation that it depends on whether relatives are referred to a specialized genetic clinic. One participant stated:

*“If the family gets referred to a specialty clinic, they get [genetic] counseling. The coroner report in [city] is provided to a portion of the family doctors with a standard letter that lists all the genetics and cardio genetics programs in [city]. As a specialty clinic, we do not know how many patients receive this recommendation (denominator) we only know who comes through our door (numerator)”* (Canada)

Most participants said that either a clinical geneticist (N=76, 63%) or cardiologist (N=60, 50%) may order postmortem genetic testing. Only 47 (39%) reported that genetic counselors are allowed to order a genetic test. In many cases, postmortem genetic testing is performed on a clinical basis (N=49, 48%), in contrast with a research-based test (N=4, 4%). A third (N=32, 31%) indicated that this depends on the case. Clinical screening is recommended to surviving relatives, either concurrent with postmortem genetic testing (N=58, 56%) or after postmortem genetic testing depending on the result (N=23, 22%). Many suggested this depends on the family, where in many cases family members are already clinically screened prior to attending the cardiogenetics clinic.

“*Generally we (my cardiology colleagues) do clinical screening of relatives first to ascertain if there is a familial phenotype to guide result interpretation and then I request molecular autopsy if we have a suspicion of a diagnosis or if nothing is found on relative*.” (Europe)

Overall 45% of participants (N=54) replied that the hospital or healthcare system pay for postmortem genetic testing. Others (N=19, 16%) reported it was covered by the healthcare insurance of the deceased or surviving family members, or that family members have to pay for it themselves (N=17, 14%).

### Follow-up of surviving relatives

64% of participants responded that certain follow-up care for surviving relatives was provided (N=78), including either clinical screening (N=75) or post-test genetic counseling (N=78). Further, 47 participants (39%) reported that psychosocial care for relatives was provided. Only four participants said that there was no follow-up in place.

### Attitudes towards practices for postmortem genetic testing

Table 2 shows survey responses regarding attitudes towards practices of postmortem genetic testing. A majority of participants considered the local approach towards postmortem genetic testing not entirely (N=55, 55%) or not at all (N=8, 8%) effective or efficient, while 37 (37%) participants replied that they indeed thought their practice was effective/efficient. Relatively more healthcare professionals from North America reported that they thought the approach was not entirely effective (70%), compared to healthcare professionals from Europe (55%) and Australia/New Zealand (45%), although this was not considered significant based on a Bonferroni-corrected *p* value (*p* = 0.015). Half of participants believed that processes used in their practice are sufficient to support postmortem genetic testing (N=50, 50%), while 32% (N=32) believed that this was sometimes the case.

**Table 2.** Attitudes of healthcare professionals towards practices regarding postmortem genetic testing

|  | Total | Europe | Australia&New Zealand | NorthAmerica |  | Other |
| --- | --- | --- | --- | --- | --- | --- |
|  | N (%) | N (%) | N (%) | N (%) | P value<sup>b</sup> | N (%) |
| **Approach is effective/efficient in the work setting<sup>a</sup>** |  |  |  |  |  |  |
| Yes | 37 (37.0) | 17 (42.5) | 15 (51.7) | 3 (13.0) | 0.015 | 2 (25.0) |
| No, not entirely | 55 (55.0) | 22 (55.0) | 13 (44.8) | 16 (69.6) |  | 4 (50.0) |
| No, not at all | 8 (8.0) | 1 (2.5) | 1 (3.4) | 4 (17.4) |  | 2 (25.0) |
| **Family members adjust/cope with postmortem genetic test results<sup>a</sup>** |  |  |  |  |  |  |
| Yes | 37 (37.0) | 17 (42.5) | 9 (31.0) | 11 (47.8) | 0.076 | 0 (0.0) |
| No | 1 (1.0) | 1 (2.5) | 0 (0.0) | 0 (0.0) |  | 0 (0.0) |
| On a case-by-case basis | 54 (54.0) | 15 (37.5) | 19 (65.5) | 12 (52.2) |  | 8 (100.0) |
| Unsure | 8 (8.0) | 7 (17.5) | 1 (3.4) | 0 (0.0) |  | 0 (0.0) |
| **Processes are sufficient to support postmortem genetic testing occurring in their clinic<sup>a</sup>** |  |  |  |  |  |  |
| Yes | 50 (51.0) | 25 (64.1) | 15 (53.6) | 10 (43.5) | 0.051 | 0 (0.0) |
| No | 16 (16.3) | 3 (7.7) | 3 (10.7) | 8 (34.8) |  | 2 (25.0) |
| Sometimes | 32 (32.7) | 11 (28.2) | 10 (35.7) | 5 (21.7) |  | 6 (75.0) |
| **Processes in place perpetuate health inequities<sup>a</sup>** |  |  |  |  |  |  |
| Yes | 50 (50.0) | 18 (45.0) | 13 (44.8) | 16 (69.6) | 0.257 | 3 (37.5) |
| No | 28 (28.0) | 13 (32.5) | 11 (37.9) | 3 (13.0) |  | 1 (12.5) |
| Unsure | 22 (22.0) | 9 (22.5) | 5 (17.2) | 4 (17.4) |  | 4 (50.0) |

<sup>a</sup> Statistical significance tests are not performed on questions with multiple answer options
<sup>b</sup> Chi square analyses were performed on continents Europe, Australia/New Zealand and North America, due to low numbers of responses from other continents

50% of participants (N=50) believed that there are processes in their clinic/country that perpetuate health inequities in care for surviving relatives and access to postmortem genetic testing, primarily due to limited coverage of insurance companies, geographical location, and lack of resources and education of coronial or specialist services. One participant explained:

*“The process is arduous for families, so families with less support and fewer resources are less likely to undergo postmortem genetic testing and the care that accompanies it*.*”* (United States)

Another participant said:

“*The system is not known. Only people aware can access and are able to find economical support for postmortem genetic testing*.” (Europe)

## DISCUSSION

Postmortem genetic testing is not routinely performed after a SCD in the young across all continents involved in this study. Indeed, this study reveals striking inconsistencies within and between countries and continents, in access to all aspects of the postmortem genetic investigation, from the completion of an autopsy, to access to clinical and genetic investigations.

Global practices are remarkably inconsistent, often operating on a chance basis according to which investigating pathologist or medical investigator team is involved. A majority of healthcare professionals reported that the procedures in place for postmortem genetic testing are largely ineffective, with healthcare professionals in North America more often indicating this compared to healthcare professionals in Europe and Australia/New Zealand. Importantly, these responses are largely from those who are more likely to have links to investigative services than those not surveyed. The true picture, including practices undertaken by non-specialists, is likely to be considerably worse. Since failure to identify genetic causes of SCD can result in further deaths within the families involved, access to specialized multidisciplinary clinics with standardized practice clearly needs to be improved.

Many healthcare professionals highlighted the dependency on the mortuary or coroner/medical examiner involved, or on the bereaved family members to collate and make available key information, often due to lack of experience or systematic procedures. As indicated by Michaud et al (18), police or coroners are often the only people who communicate with the family and they are likely to miss relevant information about the medical individual or family history. Increasing knowledge regarding the importance of postmortem genetic testing in young SCD cases seems to be pivotal for improving the procedures worldwide. Further, involvement of a genetic counselor or clinical geneticist is considered important for obtaining this data as well as for appropriate information provision to the family (18, 19). Access to genetic counseling is regularly available to only 60% according to healthcare professionals participating in this study.

Our findings indicate that financial resources to implement routine procedures are lacking for hospitals or institutes, however less so for Australia and New Zealand. This likely provides barriers for families in accessing accurate information about postmortem genetic testing and clinical surveillance of at-risk relatives. Previous studies examining practices around postmortem procedures among forensic pathologists in Europe and genetic counsellors in the United States showed financial resources were limited for the majority and postmortem samples were not regularly collected or stored (16, 19). This corresponds to our findings, reflecting a largely case-by-case approach, commonly reflecting the local knowledge or bias of the pathologist involved in the case, especially in Europe and North America. This likely perpetuates inequities whereby families with resources can navigate and find appropriate care, while others do not. Systematic processes, as well as adequate resources, are needed to ensure all families who experience a SCD can receive the recommended standard of care, ideally including autopsy, postmortem genetic testing – including genetic counselling - and clinical monitoring of surviving relatives at a multidisciplinary cardiogenetic clinic (7).

Psychological support is infrequently offered to families experiencing a SCD. A needs analysis study by McDonald et al. (20) identified that psychological needs of surviving family members were the most unmet, and a recent registry-based study showed only 12% of surviving relatives sought referral to a clinical psychologist (21). Up to 50% of family members will suffer from post-traumatic stress symptoms and prolonged grief, on average 6 years after the SCD (22). Our findings demonstrate psychosocial support is only implemented in the care for these families in a minority of settings, often on a case-by-case basis. It is important to identify how to best provide adequate psychosocial support for family members to help cope with bereavement.

While we achieved reasonable widespread international representation, this study was limited by the overall small sample size and with only a few participants from Asia, South America and Africa. As indicated, numerous networks and personal contacts of the researchers were approached to inform healthcare professionals about this study to improve the response. Due to the small sample, comparison of practices among countries and continents were limited. Given the field of cardiogenetics and postmortem genetic testing is overall small, we were fortunate to have participants with significant expertise in postmortem genetic testing allowing for unique insights. Yet this is likely to have led to a positively biased perspective, as these experienced healthcare professionals probably have more processes in place compared to healthcare professionals working in less experienced settings.

## CONCLUSION

We show that access to postmortem genetic testing is highly variable between countries, and that many aspects of the process of the investigation are largely inconsistent, leaving many cases inadequately investigated. Lack of financial resources and limited knowledge about the importance of postmortem genetic testing among professionals involved may lead to inequities in access for many SCD families. Since the genetic underpinnings of many cases of SCD are now well understood, health services should be aiming for postmortem genetic testing to be made available in all cases of sudden death in the young <40 years. To achieve this, education of all health and legal professionals involved is needed, as well as systematic and clear processes for dealing with a young SCD. These services require strong leadership and adequate resourcing. The model of the specialized multidisciplinary investigative service as described by recent international guidelines is the gold standard for management of families following a young SCD, and efforts to ensure these are accessible equitably around the world are needed (7).

## DATA AVAILABILITY

Data will be made available upon reasonable request to the corresponding author.

## Supporting information

STROBE checklist

## Data Availability

Data will be made available upon reasonable request to the corresponding author.

## ACKNOWLEDGEMENTS

We thank all the healthcare professionals who participated in this survey.

## AUTHOR INFORMATION

1. Conceptualization: LvdH, PvT, JI; 2. Data curation: LvdH; 3. Formal Analysis: LvdH, LY; 4. Funding acquisition: LvdH, JI; 5. Investigation: LvdH, PvT, JI; 6. Methodology: LH, CAJ, HM, JS, JD, CS, PT, JI; 6. Project administration: LH, JD, LY, JI; 7. Resources: PT, JI; 8. Software: Not Applicable; 9: Supervision: PvT, JI; 10. Validation: Not Applicable; 11. Visualization: LvdH; 12. Writing – original draft: LvdH; 13. Writing – review & editing: JD, LY, HM, CAJ, JD, JK, CS, PT, JI.

## ETHICS DECLARATION

Ethical approval of the local institutional ethics committee was obtained (Sydney Local Health District, RPAH Zone). This study was conducted in accordance to the principles of the Declaration of Helsinki. Completion of the survey was deemed as informed consent given. Individual-level data was de-identified.

