## Supplementary table 1 for "Global approaches to postmortem genetic testing after sudden cardiac death in the young: A survey among healthcare professionals"

**S Table 1** Professional organizations asked for recruitment of patient sample

**Patient organizations**

**American**

National Society of Genetic Counselors (NSGC)

**European**

European Reference Network of cardiogenetics (ERN)

European Society of Human Genetics (ESHG)

Dutch Working group of Cardiogenetics (VKGN)

British Society for Genetic Medicine (BSGM)

**Australian**

Australian Society of Genetic Counselors (ASGC)

Human Genetics Society of Australasia (HSGA)

Cardiovascular Genetics Flagship

**Asian**

Human Genetics Society of Australasia (HSGA)
